# Cerebral Thrombus Analysis in Infective Endocarditis: Unveiling Composition for Diagnostic Insight

**DOI:** 10.1101/2023.10.10.23296826

**Authors:** Aurora Semerano, Beatrice Dell’Acqua, Manuel Montano, Francesca Sanvito, Angela Genchi, Ghil Schwarz, Andrea Bergamaschi, Michela Sampaolo, Giorgia Serena Gullotta, Andrea Falini, Pietro Panni, Elio Clemente Agostoni, Guillaume Saliou, Steven David Hajdu, Luisa Roveri, Patrik Michel, Gianvito Martino, Massimo Filippi, Davide Strambo, Marco Bacigaluppi

## Abstract

**Background and Aims:** Infective endocarditis (IE) is a life-threatening condition known to cause stroke. Swift diagnosis and antibiotic treatment are crucial for preventing cerebral and systemic embolism, therefore reducing mortality and morbidity. However, diagnosing IE can be challenging. In this study, we aimed to assess whether analyzing cerebral thrombi retrieved by endovascular thrombectomy from stroke patients with IE could aid in the diagnosis and shed light on the composition signature of endocarditic thrombi.

**Methods:** We compared cerebral thrombi from three groups of ischemic stroke patients: those with definite infective endocarditis (IE) (n=10), those with cardioembolic stroke and concomitant infections other than IE (n=10 CE-I^+^), and those with cardioembolic stroke without infections (n=30 CE-I^-^). Our multiparameter analysis encompassed histological examinations, molecular biology and microbiological tests to detect microorganisms within the thrombi and to comprehensively assess their structural composition and immune signatures.

**Results:** We directly detected invading pathogens through histology or PCR in all cerebral thrombi from IE patients, while none of the control thrombi exhibited such pathogens. Thrombi from IE patients displayed a distinct composition, characterized by a significant lower content of red blood cells, reduced CD14+ monocytes, increased von Willebrand Factor density, and a cell-dominant pattern of Neutrophil Extracellular Traps (NETs) deposition.

**Conclusions:** Comprehensive analysis of cerebral thrombi from stroke patients with suspected IE sustains early, definitive endocarditis diagnosis by detecting pathogens and immunothrombotic changes.

## Introduction

In the last 20 years, the incidence and mortality due to Infective Endocarditis (IE) have increased by 2.3 times^1^. Despite improvements in its management, IE is still associated with a high mortality rate, that is partly related to neurological complications of the disease^2^. Ischemic stroke (IS) from embolization of vegetations complicates IE in the 20% to 40% of cases^3^, and is often one of the presenting symptoms of IE ^4^. The risk of stroke is higher at the time of IE diagnosis and decreases rapidly after the initiation of antimicrobial therapy^5^, underscoring the importance of prompt diagnosis to prevent embolization. In addition, identifying the causative microorganism and determining its sensitivity profile are crucial to optimize treatment and reduce drug toxicity^6^.

However, recognizing the signs and symptoms of IE and establishing an etiological diagnosis can be particularly challenging especially in the acute stroke setting. Indeed, up to 30% of patients with IE present with negative findings on echocardiography or blood cultures^7, 8^. Even the histological analysis on valvular specimens obtained after heart surgery, which is a cornerstone for the definite diagnosis of IE, may be not feasible because of delayed times of surgery due to the neurological conditions, or not informative due to the already ongoing empiric antimicrobial therapy.

Clinical outcome after IE has shown limited improvement over the last decades^9, 10^. This is partly due to the incomplete understanding of the precise pathological mechanisms and the complex interplay of microorganism virulence, coagulation processes, and immune response, which contribute to the formation, growth, and embolization of endocarditic vegetations. A better knowledge of these pathophysiological processes could offer novel therapeutic targets for IE^11, 12^.

The development of endovascular thrombectomy as an acute treatment for large vessel occlusion strokes, allows the analysis of retrieved cerebral thrombi, which is emerging as a relevant opportunity to complement the diagnostic investigation of stroke etiology and to provide new insights into stroke pathogenesis^13, 14^. The most recent ESC Guidelines 2023 first introduce that, when performing endovascular thrombectomy, the retrieved embolic material should undergo pathological and microbiological analyses^15^. However, the specific sample processing and histological features defining IE in cases of cerebral embolization remain unformalized.

In this study, we aimed to 1) assess the ability of histological and microbiological analyses to identify microorganisms within cerebral thrombi and evaluate their diagnostic performance in the diagnosis of IE; 2) characterize the structural composition and inflammatory infiltration of cerebral thrombi in IE.

Finally, this article identifies and summarizes pragmatic aspects of the analysis of cerebral thrombi in the setting of IE that could promote the implementation of current recommendations.

## Material and Methods

### Study population

This study was conducted on consecutive acute IS patients treated by endovascular thrombectomy at two comprehensive stroke centers (San Raffaele Hospital, Milan, Italy; Lausanne University Hospital, Lausanne, Switzerland) between April 2017 and September 2022, and with cerebral thrombi available for histological analysis. Patients’ clinical and radiological data were prospectively collected respectively in the local hospital stroke registries.

For this study we selected:

i. consecutive patients diagnosed with definite IE according to the modified Duke clinical criteria after a complete diagnostic work-up including blood cultures and transthoracic and/or transesophageal echocardiography (IE group).
ii. a control group of patients diagnosed with cardioembolic stroke according to the TOAST criteria, with no evidence of any concomitant infection at the time of stroke onset (CE-I^-^ group), matched 1:3 with IE patients, for age, sex, intravenous thrombolysis, and previous anti-thrombotic treatment.
iii. a second control group of consecutive patients in equal number to the IE group, with cardioembolic stroke according to TOAST criteria, and concomitant infection other than IE at the time of stroke onset (CE-I^+^ group). This was defined as suggestive symptoms (i.e., cough, dyspnea, pleuritic pain, urinary tract symptoms, etc.) and history of fever within the previous week of stroke onset and/or determination of body temperature >37.5°C at admission.

The collection of patients’ data and cerebral thrombi analysis was approved by local Ethics Committee at each hospital and was performed in accordance with the ethical standards laid down in the 1964 Declaration of Helsinki and its later amendments. Subjects gave informed consent for participation in the study.

### Clinical variables

For each patient we recorded demographic data, vascular risk factors, clinical history, administration of thrombolytic treatment, stroke severity assessed by the NIH Stroke Scale (NIHSS) on admission and at the discharge, imaging and procedural data (Alberta Stroke Program Early CT, ASPECTS score, presence of hyperdense vascular sign, occlusion site, type of thrombectomy device, number of procedure maneuvers, reperfusion degree by modified Treatment in Cerebral Ischemia, mTICI), stroke etiology according to TOAST classification, therapy at stroke onset, laboratory values within 24 hours from stroke symptom onset, and 3-month functional outcome assessed by the modified Rankin Scale. Presentation of IE was considered subacute if signs and symptoms had started 1 to 6 months prior to index stroke, and acute when signs and symptoms had started less than one month prior to diagnosis.

### Cerebral thrombi analysis

#### Histology

Immediately after retrieval during mechanical thrombectomy, cerebral thrombi were fixed in 10% formalin and stored at +4 °C until processing. Formalin-fixed specimens were then longitudinally embedded in paraffin and cut in serial sections of 5μm. Thrombus analysis was centrally performed at the San Raffaele Scientific Institute, Milan, Italy.

To detect the presence of bacteria or fungi, cerebral thrombi retrieved from patients from the IE group and the control groups CE-I+ were stained with hematoxylin and eosin (H&E), Grocott’s methenamine silver (GMS), GRAM and/or Periodic Acid Schiff (PAS) staining and evaluated at 63x or higher magnification by an expert pathologist^16, 17^.

Histological examination to analyze and quantify the different thrombus components included hematoxylin and eosin (H&E), Martius Scarlet Blue (MSB) staining, and Prussian blue staining. The content of Red Blood Cells (RBCs) and fibrin was assessed on MSB^18^. To quantify other thrombus components object of the study, we performed immunohistochemical staining using the following antibodies: platelets (CD61^+^ areas; anti-CD61, 1:100, Dako), neutrophils (MPO^+^ cells; anti-MPO, 1:500, Dako), macrophages (CD68^+^ cells; anti-CD68 PG-M1, 1:75, Dako), neutrophil extracellular traps (citH3^+^ areas; anti-citH3, 1:200, Abcam), T lymphocytes (CD3^+^ cells; anti-CD3, clone 2GV6, Ventana, 790–4341), B lymphocytes (CD20^+^ cells; anti-CD20, clone L26, Ventana,760–253), monocytes (CD14^+^ cells; anti-CD14, 1:100, Biorad, AHP1059), vWF (vWF^+^ areas; anti-vWF, 1:1000, Abcam, ab6994), endothelial cells (CD34^+^ cells, anti-CD34, 1:500, Invitrogen, MA1-10202), fibroblast (smooth muscle cells, SMA^+^ cells, anti-aSMA, 1:200, Sigma, A 2547).

Stained sections were scanned with Aperio® Microscope Digitizer (Leica Biosystems), at 20x magnification. Superficial areas of RBCs, fibrin, platelets, NETs and VWF were automatically quantified using the classification algorithm of Orbit Image Analysis® (v3.64) Software. Results were expressed as percentage of positive areas on the total thrombus area. Cellular elements (neutrophils, macrophages, T and B lymphocytes and monocytes) were automatically counted using the object segmentation algorithm (Mumford-Shah segmentation algorithm) of Orbit Image Analysis® (v3.64) Software, and results were expressed as cells/mm^2^.

The morphological features of NETs were visually assessed by three independent investigator blinded to clinical and radiological information, and thrombi were categorizing thrombi into cell-dominant or web-dominant pattern, as previously described^14, 19^.

Through a comprehensive evaluation of the performed stains, we also assessed the thrombus histological age based on established criteria^20^, adapted for retrieved thrombus analysis,: thrombi were classified into progressive phases, according to the prevalent pattern. Phase 1 thrombi are characterized by platelet plugging, fibrin deposition with a layered growth (Zahn’s lines), and preserved and agglomerated RBCs. In phase 2, macrophages containing hemosiderin predominate, RBCs ghosts, nuclear debris of leukocytes and fibrinous transformation are evident. In phase 3, the thrombi became hyalinized, and few leukocytes are visible between compact, fiber-rich connective tissue.

#### Molecular diagnostics

For **Polymerase chain reaction (PCR)** analysis, from paraffin embedded fragments of the thrombotic material we extracted total DNA using the QIAamp DNA FFPE Tissue Kit (Qiagen) according to the manufacturer’s instructions. DNA concentration and quality were measured. Polymerase chain reaction (PCR) amplification of the bacterial-specific 16S rDNA was performed using universal bacterial 16S rRNA primer set (16SFW 5’GATTAGATACCCTGGTAGTCCAC and 16SRW 5’ TACCTTGTTACGACTT,). *E. coli* of a pre-existing positive sample was used as a positive control and nuclease-free UV-treated water as a negative control. The PCR was performed in a ProFlex PCR System (ThermoFisher Scientific). To detect PCR inhibitions and failures in DNA extraction and to avoid false-negative results, primers bGloF and bGloR (to detect human b-globin gene) were used for each sample in the same PCR conditions. PCR results were considered valid if all controls were negative or positive as appropriate and the b-globin gene was detected in all samples. We analyzed PCR products by electrophoresis on 1.5% agarose gels. PCR products displaying a band in the expected ∼600-700 base pair region were directly sequenced with Sanger method using the same PCR primers. The samples were prepared using BigDye Terminator v.3.1 cycle sequencing kit (Applied Biosystem, Foster City, CA, USA) followed by a purification step with BigDye Xterminator™ Purification kit (Applied Biosystem, Foster City, CA, USA). We analyzed purified products on an automatic sequencer ABI PRISM 3730 genetic analyzer DNA Sequencer (Applied Biosystem, Foster City, CA, USA). The generated nucleotide sequences were analyzed with SeqScape® Software (ThermoFisher Scientific, Waltham, MA, USA) and were then compared using the BLAST program available at the National Center for Biotechnology (http://www.ncbi.nlm.nih.gov). DNA sequences were further analysed using EzBioCLoud (www.ezbiocloud.net) to provide a definitive identification of the organism to the species- or genus-level.

#### Microbiology

In two patients with IE highly suspected since hospital admission, we were able to perform a bacterial culture on a fresh fragment of the cerebral thrombus. Freshly extracted thrombi were aseptically dissected and a portion set aside for culturing. The thrombus was thus disrupted in a tissue grinder and aliquots of the prepared thrombus sediments inoculated into a BacT/Alert FA (Aerobic) and FN (Anaerobic) blood culture bottle and placed into the BacT/Alert (bioMérieux, Saint-Lauren, QC) cabinet for continuous incubation and monitor for up to 5 days^21^.

### Blood cultures

The standard procedure for adult blood culture collection at the two centers involves taking 2-3 separate sets of aerobic and anaerobic blood samples. At the local laboratories the samples are placed in the BacT/Alert system for continuous monitoring for up to 5 days. Positive blood cultures are sub-cultured on agar plates according to local standard operating procedures. The identification of these bacteria is done through a combination of physical characteristics and MALDI-TOF (Vitek MS) mass spectrometry analysis ^22^.

### Statistical analysis

Univariate tests (Mann Whitney U test for variables with two categories and Kruskall-Wallis test followed by Dunn’s post-hoc test for comparison of three groups) assessed the differences of thrombus components across subgroups. Chi-square test was used for categorical variables. P-values ::0.05 were considered statistically significant. Statistical analyses were performed with R statistical software (version3.3.2, R Core Team [2016]), and with GraphPad Prism (version 8.0).

## Results

### Clinical characteristics

During the study period we identified n=10 stroke patients undergoing EVT with cerebral thrombus available for analysis with a diagnosis of definite IE according to the modified Duke clinical criteria. A group of n=30 matched patients with cardioembolic stroke and no concomitant infection (CE-I-control group) and n=10 patients with CE stroke and non-endocarditic concomitant infection (CE-I+) were included as control groups. Baseline characteristics of the three groups are reported in **Table 1**. Presentation of IE was subacute in n=4 cases and acute in n=6. IE affected native valves in n=3 patients, valves with prosthetic ring annuloplasty in n=2 patients, bioprosthetic valve in n=2 case, mechanical prosthetic valves in n=3 cases. The mitral valve was affected in n=4, the aortic valve in n=5 cases and in n=1 case both aortic and mitral valves were involved. All patients had transthoracic echocardiograms (TTE) and/or transesophageal echocardiograms (TEE). Vegetations (n=7 cases) and valve strands (n=2) were documented on echocardiographic evaluation, periprosthetic valve thickening was reported in n=1 case, whereas perivalvular abscesses were present in n=2 cases. One patient presented also with mediastinitis. The diagnosis of IE was established in n=4 patients before and in n=6 cases after the stroke event. In n=6 cases, antimicrobial therapy was initiated before stroke onset. Three patients underwent valve replacement by open cardiac surgery after stroke. One patient with IE had also concomitant COVID-19 diagnosis at the moment of stroke onset.

**Table 1.** Characteristics of the study populations.

|  | Total (n=50) | Infective endocarditis (n=10) | Cardioembolic w/o infection (n=30) | Cardioembolic with infection (n=10) | P-value (Overall comparison) | CE-I- vs. IE | CE-I+ vs. IE |
| --- | --- | --- | --- | --- | --- | --- | --- |
| *Demographic data* |  |  |  |  |  |  |  |
| Age | 73.7 (65.9-80) | 66 (62.3-75.8) | 76.7 (70.8-81.1) | 73 (66.7-80.6) | <0.001 | 0.100 | 0.212 |
| Sex | 25 (50%) | 7 (70%) | 16 (53.3%) | 2 (20%) | 0.069 | 0.530 | 0.047 |
| *Cardiovascular risk factors* |  |  |  |  |  |  |  |
| Hypertension | 38 (76%) | 6 (60%) | 24 (80%) | 8 (80%) | 0.416 | 0.336 | 0.468 |
| Diabetes | 9 (18%) | 2 (20%) | 5 (16.7%) | 2 (20%) | 0.956 | 0.958 | 1.000 |
| Dyslipidemia | 26 (52%) | 5 (50%) | 16 (53.3%) | 5 (50%) | 0.974 | 0.975 | 1.000 |
| Coronary artery disease | 12 (24%) | 1 (10%) | 8 (26.7%) | 3 (30%) | 0.499 | 0.457 | 0.471 |
| Smoking | 6 (12%) | 2 (20%) | 2 (6.7%) | 2 (20%) | 0.364 | 0.421 | 1.000 |
| Previous stroke or TIA | 10 (20%) | 3 (30%) | 5 (16.7%) | 2 (20%) | 0.659 | 0.559 | 0.794 |
| Atrial fibrillation | 32 (64%) | 3 (30%) | 23 (76.7%) | 6 (60%) | 0.028 | 0.014 | 0.244 |
| *Antithrombotic therapy at stroke onset* |  |  |  |  |  |  |  |
| None | 17 (34%) | 1 (10%) | 11 (36.7%) | 5 (50%) | 0.149 | 0.209 | 0.108 |
| Antiplatelets | 11 (22%) | 4 (40%) | 4 (13.3%) | 3 (30%) | 0.167 | 0.139 | 0.796 |
| Anticoagulants | 22 (44%) | 5 (50%) | 15 (50%) | 2 (20%) | 0.232 | 1.000 | 0.294 |
| *Acute stroke treatment* |  |  |  |  |  |  |  |
| Intravenous thrombolysis | 3 (6%) | 0 (0%) | 0 (0%) | 3 (30%) | 0.002 | 1.000 | 0.005 |
| Onset-to-groin delay (minutes) | 255 (157-376.7) | 241 (167.9-295.8) | 255 (135-440) | 241 (195.1-298.8) | 1 | 0.614 | 0.999 |
| Baseline ASPECTS | 8.5 (8-10) | 7.5 (6-10) | 8 (8-9.6) | 10 (7.8-10) | <0.001 | 0.245 | 0.098 |
| *Occlusion site* |  |  |  |  |  |  |  |
| ICA | 10 (20.8%) | 2 (20%) | 6 (21.4%) | 2 (20%) | 0.993 | 0.993 | 1.000 |
| MCA | 33 (68.8%) | 8 (80%) | 18 (64.3%) | 7 (70%) | 0.652 | 0.556 | 0.842 |
| BA/PCA | 5 (10.4%) | 0 (0%) | 4 (14.3%) | 1 (10%) | 0.446 | 0.345 | 0.678 |
| *Collaterals* |  |  |  |  |  |  |  |
| Poor | 8 (18.6%) | 3 (33.3%) | 4 (16%) | 1 (11.1%) | 0.42 | 0.413 | 0.377 |
| Intermediate | 25 (58.1%) | 6 (66.7%) | 14 (56%) | 5 (55.6%) | 0.843 | 0.800 | 0.847 |
| Good | 10 (23.3%) | 0 (0%) | 7 (28%) | 3 (33.3%) | 0.169 | 0.158 | 0.167 |
| *EVT device* |  |  |  |  |  |  |  |
| Stent retriever alone | 7 (14.3%) | 3 (30%) | 4 (13.8%) | 0 (0%) | 0.158 | 0.333 | 0.102 |
| Aspiration alone | 27 (55.1%) | 2 (20%) | 18 (62.1%) | 7 (70%) | 0.04 | 0.037 | 0.042 |
| Stent retrieve and aspiration | 15 (30.6%) | 5 (50%) | 7 (24.1%) | 3 (30%) | 0.31 | 0.223 | 0.513 |
| *mTICI* |  |  |  |  |  |  |  |
| 2a | 4 (8%) | 2 (20%) | 2 (6.7%) | 0 (0%) | 0.235 | 0.297 | 0.177 |
| 2c | 1 (2%) | 0 (0%) | 1 (3.3%) | 0 (0%) | 0.712 | 0.734 | 1.000 |
| 3 | 45 (90%) | 8 (80%) | 27 (90%) | 10 (100%) | 0.329 | 0.551 | 0.238 |
| *Outcome measures* |  |  |  |  |  |  |  |
| mRS pre-stroke | 0 (0-2) | 0.5 (0-2.1) | 0 (0-1) | 0.5 (0-3) | <0.001 | 0.427 | 0.914 |
| Baseline NIHSS | 15.5 (8.9-22) | 16.5 (14.8-22.1) | 16 (8-22.1) | 12 (9.8-21) | 1 | 0.945 | 0.611 |
| 24-hour NIHSS | 8 (3-18) | 11.5 (3-17.1) | 7 (1-18.1) | 11 (4.3-21.3) | 1 | 1.000 | 0.985 |
| NIHSS at discharge | 4 (1-14.7) | 4 (1-15.3) | 3.5 (0-14.2) | 4 (2-16.4) | 1 | 0.997 | 0.969 |
| 3-month mRS 3-6 | 3 (1-4.2) | 3 (1-6) | 2 (1-4) | 4 (2.7-4.7) | <0.001 | 0.265 | 1.000 |
| *Blood test parameters* |  |  |  |  |  |  |  |
| Delay onset to blood sample | 22.7 (12.4-28.8) | 22.5 (10.8-24) | 19.7 (12.5-29.3) | 22.8 (16.7-35.8) | 1 | 0.785 | 0.723 |
| Total WBCs ($\times 10^3/\mu\text{L}$) | 9.2 (7.4-11.6) | 11.4 (8.9-15.1) | 8.3 (6.7-10.7) | 11.1 (8.3-13.2) | <0.001 | 0.010 | 0.630 |
| Neutrophils ($\times 10^3/\mu\text{L}$) | 6.8 (5.5-9.1) | 7.4 (6.3-10.2) | 6.3 (4.3-8.2) | 8.7 (6.4-10.6) | <0.001 | 0.213 | 0.698 |
| Lymphocytes ($\times 10^3/\mu\text{L}$) | 1.4 (1.1-1.8) | 1.2 (1-1.6) | 1.4 (1.1-1.8) | 1.4 (0.9-1.8) | 1 | 0.556 | 0.921 |
| Monocytes ($\times 10^3/\mu\text{L}$) | 0.7 (0.6-0.9) | 0.7 (0.6-0.9) | 0.7 (0.6-0.8) | 0.7 (0.5-1.1) | 1 | 0.940 | 0.787 |
| Platelets ($\times 10^3/\mu\text{L}$) | 187 (140-260) | 185 (114-272) | 183 (148-218) | 210 (125-377) | 1 | 0.875 | 0.395 |
| CRP (mg/L) | 18.1 (8.1-81.6) | 108.6 (70.7-171.9) | 9.8 (6.4-20.9) | 55.5 (15.3-75.8) | <0.001 | 0.000 | 0.012 |

Abbreviations –ASPECTS, Alberta Stroke Program Early CT Score; BA, basilar artery; CRP, C reactive protein; EVT, endovascular thrombectomy, ICA, internal carotid artery; MCA, middle cerebral artery; mRS, modified Rankin Scale; NIHSS, National Institutes of Health Stroke Scale; PCA, posterior cerebral artery; TIA, transient ischemic attack; mTICI, modified treatment in cerebral ischemia; TOAST, Trial of ORG 10172 in Acute Stroke Treatment; VA, vertebral artery; WBCs, white blood cells. Numerical variables expressed as median (interquartile range), categorical variables expressed as numer (percentage).

### Detecting the pathogen on thrombus specimen

Histopathological stain with H&E and/or GMS allowed to detect the presence of microorganisms in all the thrombi in the IE group, and in none of the CE-I^+^ control group. In most of IE thrombi (n=9/10), we observed cocci arranged in clusters and chains that could also be visualized on GMS or on GRAM staining. In the remaining IE case (1/10), GMS and PAS staining allowed instead the detection of clusters of yeast forms and pseudo hyphae with a diagnosis of fungal endocarditis (**Figure 1 A**-**C**).

**Figure 1.**
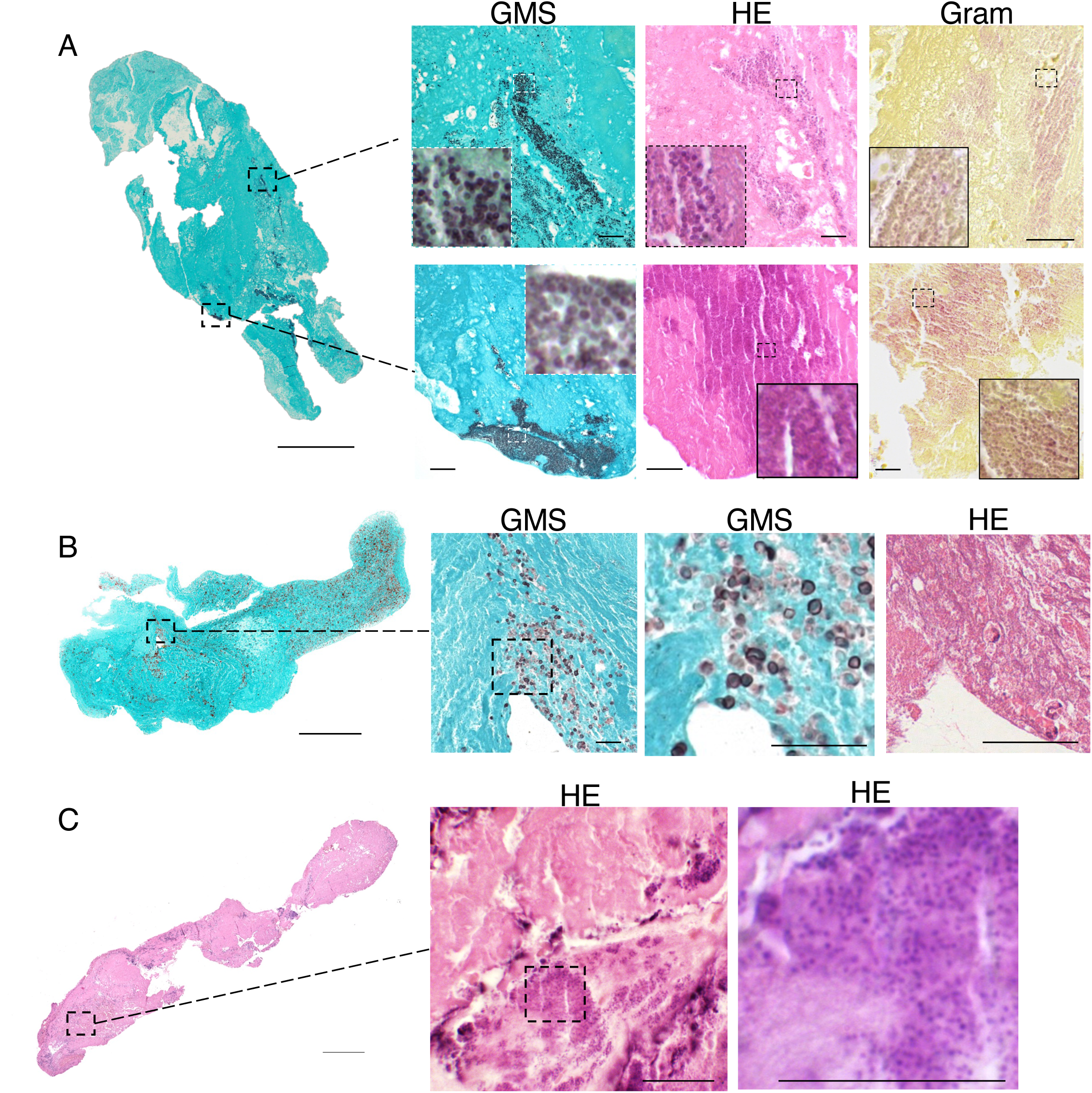
Microorganism detection on cerebral thrombi retrieved by endovascular thrombectomy in patients with IE. **A)** Low magnification view of a cerebral thrombus of a patient with bacterial IE, stained by GMS. Dark black signal at low magnification corresponds to sites of bacterial localization, as shown on the right in the higher magnification images after staining by GMS (dark black spots), Hematoxylin and Eosin (HE, dark pink spots) and Gram staining (pink spots). Scale bars: 600 µm and 30µm. **B)** Low magnification view of a cerebral thrombus of a patient with fungal IE, stained by GMS. In higher magnification of the boxed area, on the right, yeast forms can be identified on GMS staining (dark spots) and corresponding H&E staining. Scale bars: 600 µm and 30µm. **C**.) Low magnification view of a cerebral thrombus of a patient with bacterial IE after H&E staining. While on low magnification bacterial colonies cannot be promptly detected, in the higher magnification on the right the colonies (in dark pink) become clearly visible. Scale bars: 600 µm and 30µm.

PCR analysis on the thrombotic material was performed for n= 7 out of 10 patients in the IE group and for n=6 out of 10 patients in the CE-I^+^ control group. For the remaining patients in both IE and CE-I^+^ groups, PCR analysis was not possible due to insufficient quantity of material.

Among the IE thrombi tested (n=7), Streptococcus spp were detected in 3 cases, in one case PCR analysis yielded a positive result for Pseudomonas spp, while in 3 cases no specific pathogen could be detected by PCR. In the CE-I^+^ control group none of the 6 thrombi showed the presence of IE-related microorganism. The detection of Cutibacterium acnes in a single case was considered as contamination.

The bacterial culture performed on fresh fragments of the cerebral thrombus in two patients resulted in the growth of Streptococcus gallolyticus in one case, and Staphylococcus epidermidis in the other. The latter result was deemed indicative of contamination due to the rare incidence of Staphylococcus epidermidis IE on native valves, its higher likelihood of being found as a contaminant in biological samples, and the concomitant blood cultures positive for Streptococcus gallolyticus, a well-known agent of IE on native valves.

The details of microorganisms detected on PCR analysis on cerebral thrombi, thrombus cultures and blood cultures in each IE patient are shown in **Table 2**. In one case (#06), thrombus PCR were concordant with thrombus culture (Streptococcus gallolyticus), while blood culture resulted negative, supporting the initiation of antimicrobial therapy; in another case (#07), thrombus PCR revealed a Streptococcus spp but actual blood cultures were positive for Staphylococcus aureus in a patient known for a previous, clinically-resolved streptococcal endocarditis eight months earlier, suggesting the possibility of residual DNA material from the previous infection that could explain the PCR result^23^. Further, in one thrombus (#04) a Pseudomonas spp was found on PCR amplification in a patient with known blood cultures positive for Staphylococcus; this finding, along with the detection of cocci on GMS histological staining, was interpreted as a possible non-specific amplification, consequence of manipulation passages of the sample during pre-analytic phases.

**Table 2.** Compared results of sequencing of PCR positive thrombus section to histopathology, thrombus, and blood culture.

| Thrombus | Valve type | Blood cultures | Valve |  | Thrombus |  |  |
| --- | --- | --- | --- | --- | --- | --- | --- |
|  |  |  | Valve histology | Valve cultures | Thrombus histology | Thrombus PCR | Thrombus culture |
| #01 | Aortic bioprosthetic valve | Candida parapsylosis | np | np | Positive (Hypae) | np | np |
| #02 | Native mitral valve with prosthetic ring anuloplasty | Staphylococcus aureus | Positive (cocci) | Negative | Positive (Cocci) | np | np |
| #03 | Mechanical prosthetic aortic valve | Streptococcus dysgalactiae | np | np | Positive (Cocci) | np | np |
| #04 | Native aortic valve | Streptococcus mitis | np | np | Positive (Cocci) | Pseudomonas spp | np |
| #05 | Mechanical prosthetic aortic valve | Streptococcus gordonii | np | np | Positive (Cocci) | Streptococcus spp | np |
| #06 | Native mitral valve with prosthetic ring anuloplasty | Negative | np | np | Positive (Cocci) | Streptococcus spp | Streptococcus gallolyticus |
| #07 | Mechanical prosthetic mitral valve | Staphylococcus aureus | np | np | Positive (Cocci) | Streptococcus spp | np |
| #08 | Mechanical prosthetic aortic valve | Streptococcus epidermidis | np | Negative | Positive (Cocci) | Negative | np |
| #09 | Native mitral and aortic valves | Enterococcus faecalis | np | np | Positive (Cocci) | Negative | np |
| #10 | Native mitral valve | Streptococcus gallolyticus | np | np | Positive (cocci) | Negative | Staphylococcus epidermidis |

Abbreviations – MSSA, Meticillin-Sensitive Staphylococcus aureus; np, not performed/not available.

### Structure, immune phenotype, and age of cerebral thrombi of stroke patients with endocarditis

For the analysis of clot composition, we excluded the sample from a patient in the IE group who had concomitant COVID-19, due to the potential alteration of the structural and immune cell composition of the thrombus associated to COVID-19^24^.

Endocarditic thrombi had markedly reduced RBCs content compared to control groups (% over total thrombus area: mean ±SE= 44.58±4.60, 50.78±4.73, 13.97±4.05 in CE-I^-^, CE-I^+^ and IE respectively, p≤0.001, **Fig. 2A**) and significantly higher von Willebrand factor (vWF) density (% over total thrombus area: mean ±SE= 14.90±1.45, 15.65±1.97 and 24.23± 2.61 in CE-I^-^, CE-I^+^ and IE, respectively, p≤0.001; **Fig. 2C**). The fibrin and platelet content was similar in the three groups (fibrin % over total thrombus area, on MSB staining^25^: mean± SE= 37.50±5.08, 26.92.1±6.29, 41.98±12.39 in CE-I^-^, CE-I^+^ and IE respectively, p>0.05; **Fig. 2A**; platelet % over total thrombus area: mean± SE= 25.72± 2.82, 33.17±5.627 and 25.41± 2.36, in CE-I^-^, CE-I^+^ and IE, respectively, p≥0.05; **Fig. 2B**).

**Figure 2.**
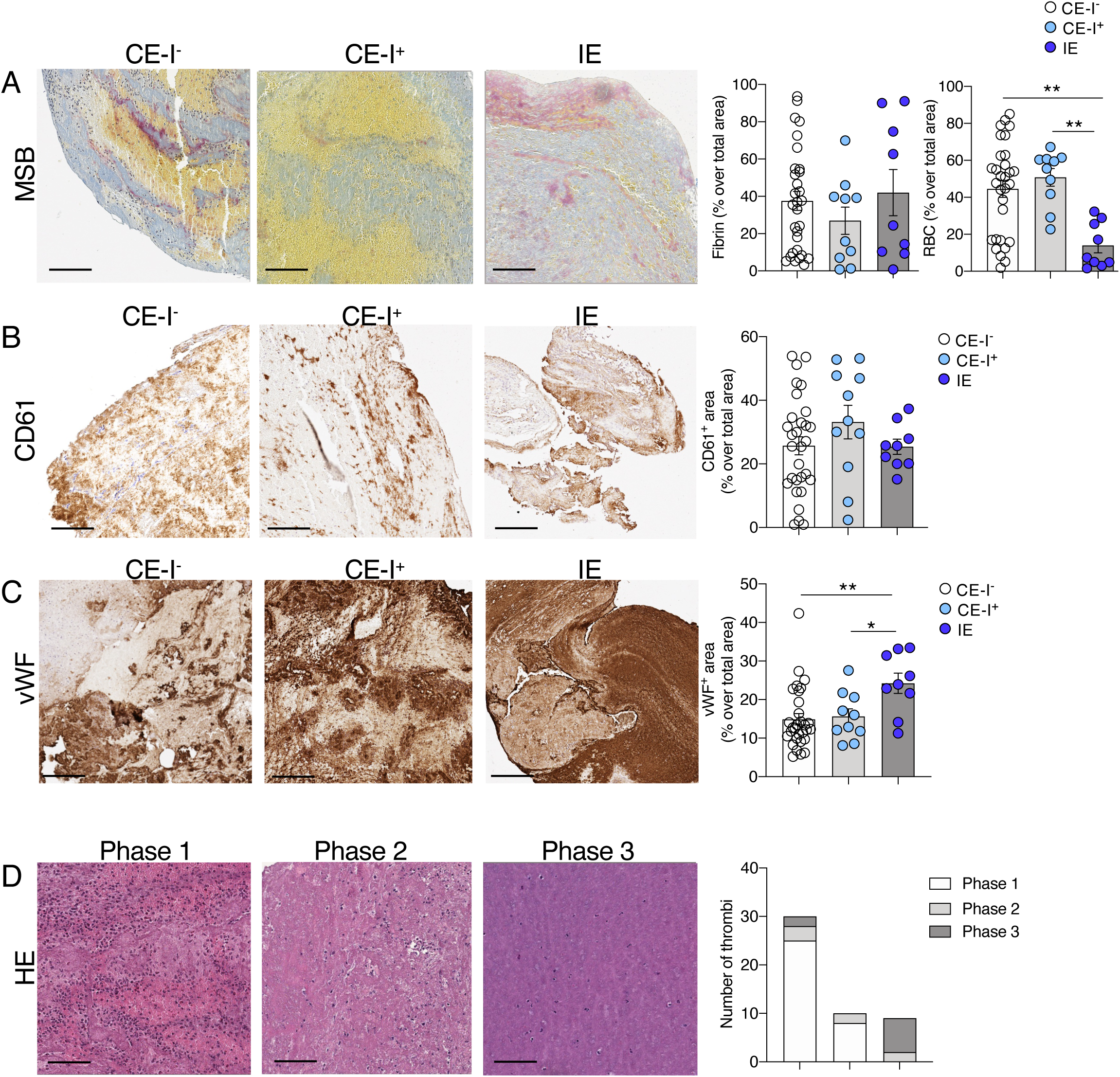
Structural composition of cerebral thrombi of patients with IE and controls. **A)** Quantification and representative images of fibrin (pink component) and RBCs (yellow component) in cerebral thrombi of CE-I^-^ (n=30), CE-I^+^ (n=10) and IE (n=9) patients, stained with MSB. Kruskall-Wallis test, Dunn’s post-hoc test, **p≤0.01. **B** and **C**) Quantification and representative images of the platelet density (CD61^+^) in **B**) and von Willebrand Factor (vWF^+^) in **C**) in cerebral thrombi of patients as in A. Kruskall-Wallis test, Dunn’s post-hoc test, *p≤0.05, **p≤0.01. **D**) Representative H&E images of cerebral thrombi in different aging-phases (from Phase 1 to Phase 3) and distribution of the age-phases of the thrombus depending on stroke etiology (CE-I^-^, CE-I^+^, IE). Chi-square, ****p≤0.0001. Scale bars in A, B, D: 100 µm, in C: 200µm.

The inflammatory cell analysis revealed that IE thrombi had a significant lower density of infiltrating monocytes (**Fig. 3A**; 1042.0 ±109.4,1410.0 ±258.4 and 470.5±99.6, CD14^+^ cells per mm^2^, in CE-I^-^, CE-I^+^ and IE, respectively, p≤0.001). However, IE thrombi did not differ in terms of macrophages (CD68^+^ cells), T-Cells (CD3^+^ cells), B-Cells (CD20^+^ cells) and neutrophils (MPO^+^ cells) (**Fig. 3B**-**D**). The density of neutrophil extracellular traps (NETs) did not significantly change in thrombi from the different groups as well as the normalized density of NETs released by neutrophils (NET-to-MPO ratio) (**Fig. 3E**). However, the pattern of NETs was different: endocarditic thrombi had a prevalent cell-dominant NET pattern while the control groups were rather characterised by a web-dominant NET pattern (**Fig. 3F**). Considering the relative inflammatory cell infiltrate in endocarditic thrombi, neutrophils were the rather prevalent cell type that could be found (**Fig. 3G**).

**Figure 3.**
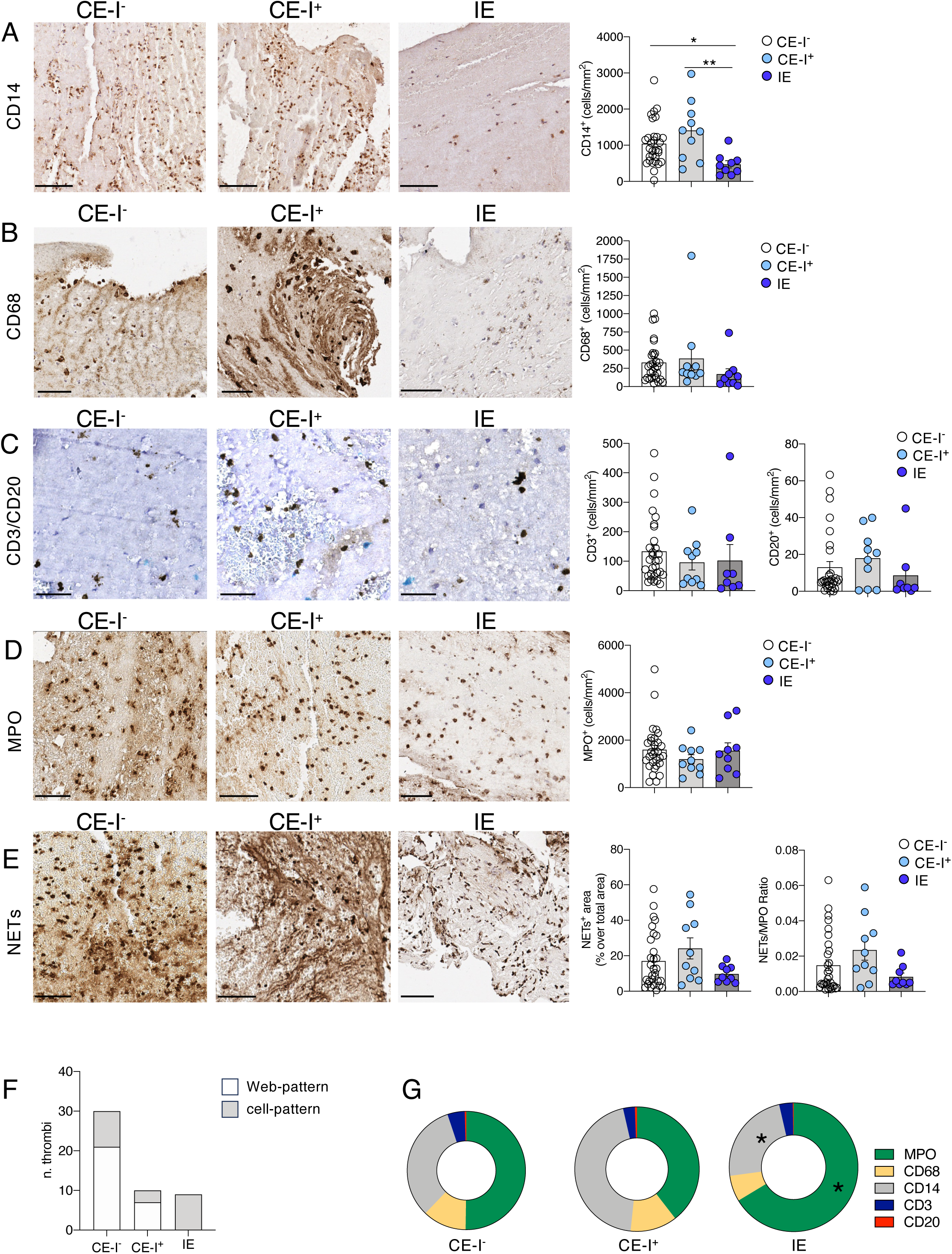
Immune cell composition of cerebral thrombi of patients with IE and controls. **(A-E)** Representative images and quantification of monocytes (CD14^+^ cells) in **A**), macrophages (CD68^+^ cells) in **B**), T- and B-lymphocytes (CD3^+^ cells in brown and CD20^+^ cells in blue) in **C**), neutrophils (MPO^+^ cells) in **D**) and neutrophils extracellular traps (NETs, citH3^+^) in **E**) in cerebral thrombi of CE-I^-^ (n=30), CE-I^+^ (n=10) and IE (n=9) patients. Shown is also the normalized NET-to-MPO ratio in **E**). Kruskall-Wallis test, Dunn’s, *p≤0.05, **p≤0.01. **F)** Distribution of the NET pattern (cell pattern and web-pattern) across thrombi of different etiology as in A. Chi-square test, ****p=0.0005. Scale bars in A, B: 80 µm, in C: 40µm, in D, E: 60 µm. **G)** Percentage of inflammatory cell subtype according to stroke etiology, CE-I^-^, CE-I^+^ and IE. Chi-square, *p≤0.05.

The assessment of thrombus histological age revealed that in the two control groups thrombi were mainly of recent formation with most thrombi classified into phase 1; conversely, thrombi from endocarditic stroke patients were older as they were significantly more often classified into phase 3 (p≤0.0001; **Fig. 2D**). Accordingly, endocarditic thrombi displayed features for endothelium (anti-CD34), fibroblasts (anti-SMA), and hemosiderin deposits (blue on Prussian blue staining), all hallmarks of thrombi with an age at least older than 10 days^20^, data not shown.

## Discussion

In this study on patients with stroke and IE, we found that: i) histopathological stains successfully identified microorganisms within cerebral thrombi from IE; ii) bacterial PCR and culture of cerebral thrombi provided valuable supplementary diagnostic information in selected cases; iii) cerebral thrombi from IE patients exhibited distinctive features, including hallmarks of more advanced age, lower red blood cell and CD14+ cell densities, increased von Willebrand factor (vWF) content, and a higher prevalence of NETs with a cell-dominant pattern.

While some previous case reports and small case series already described the results of thrombus analysis in IE patients, with findings in line with ours^26–33^, this is the first study to systematically evaluate the contribution of multi-parametric thrombus analysis to the diagnosis of infective endocarditis. The three methods we employed to detect pathogens on cerebral thrombi, including histological analysis, PCR, and thrombus culture, have been previously adopted in studies on cardiac valves^34^. On cardiac valves, histopathology demonstrated 100% specificity for IE and showed good sensitivity ^21^. In our study, histological analysis of cerebral thrombi demonstrated comparable diagnostic performance, with a very high sensitivity and specificity, although these findings should be interpreted cautiously due to the limited sample size. Crucially, the direct visualization of bacteria or fungi in thrombus specimens can strongly suggest an IE diagnosis prompting further clinical and microbiological analysis and supporting the initiation of empiric antibiotic therapy. It is important to emphasize that the effective application of this technique relies on the expertise of a pathologist who carefully examines the specimens and employs various histological staining methods to confirm the presence of pathogens. This technique allows for direct visualization of pathogens, but it has the limitation of not providing classification or identification of the exact responsible microorganism. Organism-specific immunohistochemical stains, when available, can be an additional useful tool for refining pathogen identification ^35^

The second detection method, 16S rDNA PCR/sequencing, showed good sensitivity in detecting the bacterial pathogen responsible for IE on cardiac valves^22, 23^ and we observed similar results when applying this method to cerebral thrombi. In our analysis, PCR/sequencing showed lower sensitivity and specificity compared to histopathological analysis. However, this technique has the advantage of characterizing the microorganism, which provides valuable information for targeting antibiotic therapy. The results of PCR/sequencing should be interpreted with caution due to the possibility of sampling contamination during pre-analytic phases, leading to false positive results. Additionally, in patients with a history of treated IE, bacterial DNA may persist, potentially leading to discordant results or even to the idea of persisting bacteria. This could explain the two out of four patients with positive thrombus PCR but with bacterial DNA that did not correspond to the results of blood cultures. In our case series, additional analysis with thrombus culture was possible in 2 cases due to prompt clinical suspicion of endocarditis during the angiography procedure, and in one case it yielded a positive result with the identification of the sensitivity profile of IE-related microorganism. It is worth noting that studies on valve culture in infective endocarditis have reported relatively low sensitivity, ranging from 13% to 25% ^36, 37^. The administration of antibiotic therapy prior to the cultural examination plays a significant role in explaining these findings, as the sensitivity of growth on valve culture has been inversely correlated with the duration of antibiotic treatment. Prolonged antibiotic treatment reduces the likelihood of obtaining positive valve cultures^21, 38^.

In summary, a multi-parametric pathologic assessment of cerebral thrombi through histological analysis, PCR/sequencing and culture, provides valuable information for confirming the diagnosis and managing antibiotic therapy. Therefore, whenever embolized material is available in patients with IE, these methods should be considered as valuable additions to the diagnostic process. Our study contributes to the ongoing efforts aimed at improving the dissemination, implementation, and adherence to existing guidelines to align clinical practice with recommended standards^39^.

Based on our case series, we propose a flowchart to guide the process in the angiographic suite after the removal of the occluding thrombus, providing pragmatic indications for real-life implementation of guideline recommendations in order to streamline patient management pathway. When there is a high suspicion of endocarditis, the collected material should be promptly sent to the microbiology department in a sterile tube for histopathological analysis and culture of the thrombus. Conversely, in cases with no or low suspicion of underlying endocarditis, the thrombus specimen should be fixed and stored for potential delayed analysis in case doubts regarding endocarditis arise later on (**Fig. 4**). In addition to the presence of bacteria, thrombi from IE patients exhibited distinct characteristics in term of clot composition when compared to control samples, that may be attributed to the mechanisms of thrombus formation and inflammation, which have been partly studied in models of Staphylococcus-induced endocarditis^12^. Thrombus formation in endocarditis may initiate from an underlying endothelial inflammation, leading to an increased release of vWF. Indeed, vWF was abundant in induced endocarditis lesions and proved to be crucial in S. aureus adhesion to both damaged and inflamed hearth valves in mice^12^. The lower content of RBCs in IE compared to cardioembolic thrombi could in part be attributed to the age disparity of the thrombi, as endocarditic thrombi were found to be older, and it is possible that RBC degradation occurs over time^25^. The finding of predominant cellular-like pattern of NETs deposition in IE thrombi might rely on a diverse modality of secretion ^40^, or on an early bacteria-related digestion of secreted NETs to escape from the host’s defense mechanisms ^41^.While future studies are needed to better clarify the role of NETs in IE, our finding further supports NETs as integral players of immunothrombosis, due to their complex interplay with both pathogens and components of the coagulation system.

**Figure 4.**
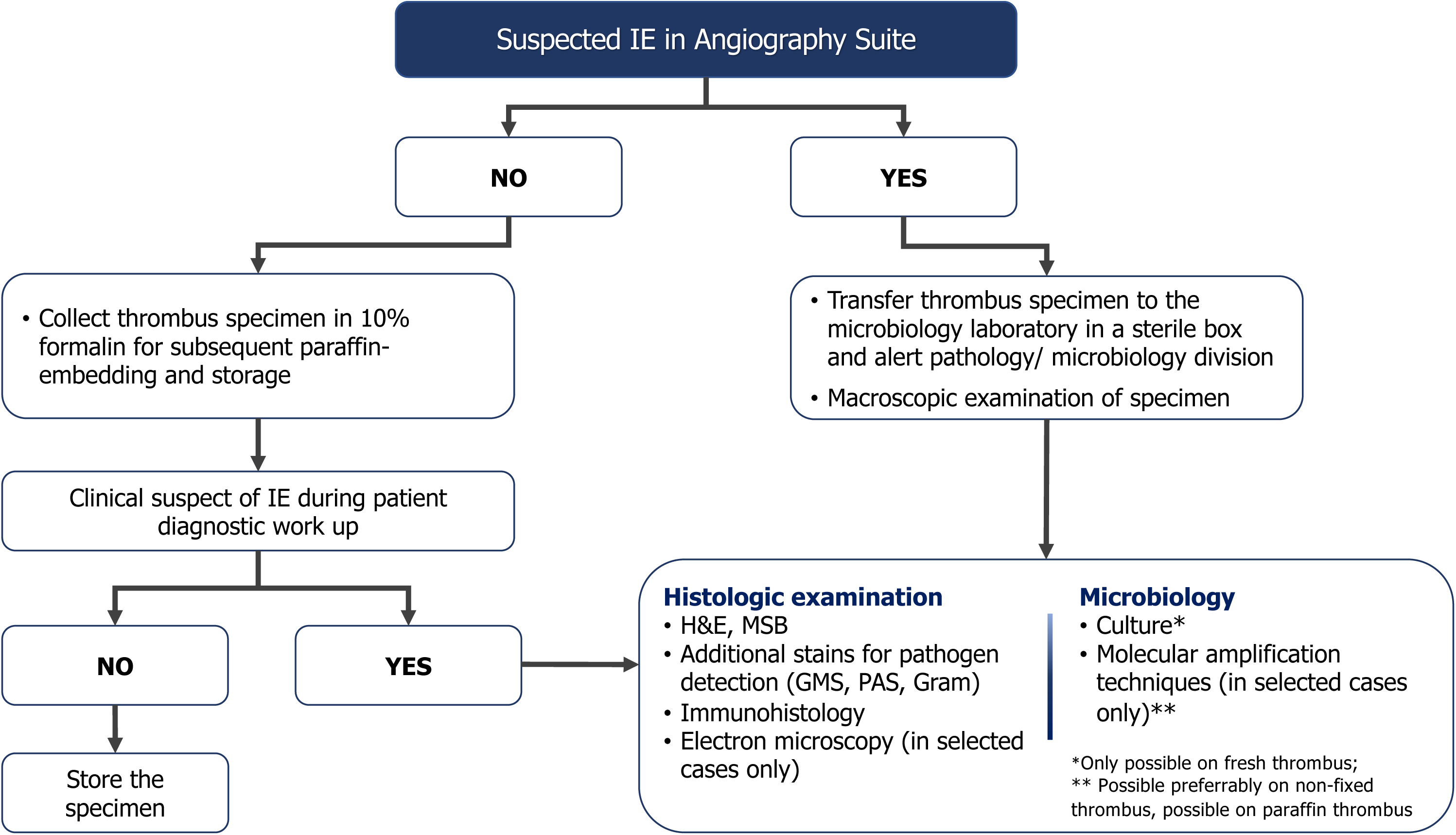
Proposed flowchart for handling retrieved cerebral thrombi with suspected infective endocarditis.

This study has certain limitations that should be acknowledged. First, the relatively small number of analyzed endocarditis thrombi may limit the generalizability of the findings. Additionally, we could not perform a direct comparison between cerebral thrombi and valve histology, which could have provided further insights. In conclusion, we propose a consistent approach in suspected endocarditis cases where the cerebral thrombus undergoes thorough processing for histological and microbiological analyses.

## Data Availability

All data produced in the present study are available upon reasonable request to the authors

## Availability of data and material

The data that support the findings of this study are available, upon reasonable request.

## Author contributions

MB was the principal investigator. Statistical analysis: AS, DS. Study concept and design: MB, AS, and DS. Acquisition of data, stainings, analysis, and or interpretation of data: AG, BDA, MM, AS, GSG, DS, GS, MS, AF, PP, ECA, GS, SDH, LR, PM, GM, MF. Drafting of the manuscript and or figure: AS, DS, MB, MF, GM.

## Disclosures

The authors declare that they have no conflict of interest.

## Funding

Dr. Strambo received research grant from the University of Lausanne and the Swiss Heart Foundation.

### Acknowledgments

We thank Elena Brambilla, Erica Butti, Stefano Grassi, for advice and support on histology. Amleto Fiocchi of the OMC for technical support and the ALEMBIC imaging facility.

## Notes

### Competing Interest Statement

The authors have declared no competing interest.

### Author Declarations

Ethics committee/IRB of San Raffaele Hospital gave ethical approval for this work

